# Identifying functional genes and pathways towards a unifying model for atrial fibrillation

**DOI:** 10.1101/2021.09.20.21263861

**Authors:** Sojin Youn Wass, Erik J. Offerman, Han Sun, Jeffrey Hsu, Julie H. Rennison, Catherine C. Cantlay, Meghan L. McHale, A. Marc Gillinov, Christine Moravec, Jonathan D. Smith, David R. Van Wagoner, John Barnard, Mina K. Chung

## Abstract

**Rationale:** Genome wide association studies (GWAS) have associated >100 genetic loci with atrial fibrillation (AF), yet the biological pathways of AF remain elusive.

**Objective:** To determine candidate causal genes associated with AF risk loci and their coexpression partners, modules, biologic and mechanistic pathways.

**Methods and Results:** Cis-expression quantitative trait loci (eQTLs) were identified for candidate genes near AF risk single nucleotide polymorphisms (SNPs) in human left atrial tissues. Genes were categorized into 3 sets according to likelihood of being a causative AF gene: 1) All Candidate Genes (with significant eQTLs or previously prioritized); 2) Any eQTL Genes (with ≥1 significant eQTL); and 3) Top GWAS SNP eQTL Genes (top SNP within the top 10 eQTL SNPs). Coexpression partners were identified for each candidate gene. Weighted gene coexpression network analysis (WGCNA) identified modules and modules with overrepresentation of candidate AF genes. Ingenuity Pathway Analysis (IPA) was applied to the coexpression partners of each candidate gene, and IPA and gene set enrichment analysis (GSEA) to each WGCNA module. 166 AF-risk SNPs were located in 135 distinct loci. The All Candidate Genes group contained 233, the Any eQTL Genes group 131 (83 novel), and the Top GWAS SNP eQTL Genes group 37 genes. IPA identified mitochondrial dysfunction, oxidative stress, epithelial adherens junction signaling, and sirtuin signaling as the most frequent pathways. WGCNA characterized 64 modules; candidate AF genes were overrepresented in 8. Modules were represented by cell injury, death, stress, developmental, metabolic/mitochondrial, transcription/translation, and immune activation/inflammation regulatory pathways.

**Conclusions:** AF candidate gene coexpression analyses suggest significant roles for cellular stress and remodeling in AF. We propose a dual risk model for AF: Genetic susceptibility to AF may not manifest until later in life, when cellular stressors overwhelm adaptive responses. These analyses provide a resource for further functional studies on potential causal AF genes.

## INTRODUCTION

Atrial fibrillation (AF) affects over 33.5 million worldwide, is strongly associated with aging and obesity, and is projected to increase further with the aging population and increasing prevalence of obesity^1^. AF is often initiated by triggers from the pulmonary vein ostia, which are targets for AF ablation, and maintained by vulnerable atrial substrates that promote AF propagation and persistence^2^. Atrial substrates become vulnerable from structural, electrical, and autonomic remodeling promoted by conditions that can cause stretch, inflammation, oxidative stress, or calcium overload^2^.

Genome-wide association studies (GWAS) have identified multiple loci associated with AF risk; the top locus is on chromosome 4q25 near *PITX2*, which has been implicated in pulmonary vein formation^3–6^. Using a collection of human left atrial appendage (LAA) tissue samples, we performed RNA sequencing (RNASeq) to identify cis-expression quantitative trait loci (cis-eQTLs), where AF GWAS SNP genotypes correlated with the expression of nearby genes; we identified candidate genes and top eQTL SNPs for 12 of 23 AF GWAS loci known at that time^3^. Since then, larger GWAS meta-analyses have identified over 100 AF risk loci, most of which have not yet undergone functional analyses^5, 6^.

Although some of the identified candidate genes have transcriptional products with known function, the biological function of many remain poorly understood in the heart. Whole-genome coexpression and pathway analysis of the candidate gene(s) associated with a target locus can provide insights into their mechanisms and regulation that can guide further functional studies^7^.

To this end, we aimed to: 1) determine candidate causal genes associated with the expanded list of AF risk loci by identification of cis-eQTLs in human LAA tissues; 2) identify coexpression partners of AF candidate genes; and 3) identify biological pathways by which these genes might function, focusing on candidate genes whose functional connections to AF are poorly understood.

## METHODS

### Left atrial tissue collection and processing

Human LAA tissues were obtained from 265 patients, including 251 who underwent elective cardiac surgery and 14 from non-failing unused transplant donors. Prior to 2008 verbal consent was obtained and documented in a process approved by the Cleveland Clinic Institutional Review Board. After this time written informed consent was obtained. Consent to use donor hearts was obtained from the family. The patient population, tissue processing, genomic DNA isolation, single nucleotide polymorphism (SNP) genotyping, RNA isolation and sequencing have been previously reported^3^. Briefly, DNA was genotyped using Illumina Hap550v3 and Hap610-quad SNP beadchips and SNP data imputed to the 1000 Genomes Project phase 2^3^. RNA unstranded 100-bp paired-end sequencing was performed on the Illumina HiSeq 2000 platform. Gene read counts are available in the Gene Expression Omnibus database (GSE69890).

### *Cis*-eQTL analyses

RNASeq and eQTL statistical analyses were previously described, with cis-eQTLs defined as being ±250 kb from the SNP^3^. Expression values were obtained from aligned files using HTseq^8^. Reads were quantile normalized, and gene counts for eQTL analysis were variance-stabilized transformed using the R package Deseq2. Samples were analyzed in R (version 3.4.4) using the surrogate variable analysis (SVA) and Limma R packages. eQTL analyses were performed for 235 samples of European descent and can be found at http://afeqtls.lerner.ccf.org3.

### Determination of candidate AF risk genes

Using our mRNA-Seq eQTL database, we determined candidate genes near the top SNPs at each locus published from two recent GWAS meta-analyses^5, 6^. Genes were categorized into 3 sets according to likelihood of being a causative AF gene, from weakest to strongest: 1) All potential candidate AF genes (<u>All Candidate Genes</u>), consisting of genes with significant cis-eQTLs (q value <0.05) from the two GWAS meta-analyses, or if no significant eQTL was identified, then the prioritized genes by Nielsen, et al^6^; 2) Genes with at least one AF SNP associated with its gene expression with q value <0.05 from our eQTL study (<u>Any eQTL Genes</u>); and 3) eQTL genes with the top GWAS SNP within the top 10 eQTL SNPs (<u>Top GWAS SNP eQTL Genes</u>).

All Candidate Genes group is the most liberal and comprehensive list, including all cis-eQTL genes and non-cis-eQTL genes prioritized by Nielsen et al. (including prioritized genes that were not expressed in our LA mRNA-Seq database). This list allows for inspection of potential AF genes, such as *PITX2* or other developmentally active genes that are no longer highly expressed in adult left atrial appendage tissue, but which are still likely to play a significant role in AF development. Any eQTL Genes group includes those showing evidence of a gene expression relationship with an AF risk SNP. Top AF GWAS SNP eQTL Genes group is the most stringent list in level of specificity, but is also the lowest in sensitivity, including genes with the highest level of evidence for future functional studies.

### Gene coexpression analyses

Coexpression analysis was performed for all genes in the All Candidate Genes group across the Ensembl genome database version 71. Each gene was included in individual linear models as a predictor with all other Ensembl genes as outcome, additionally controlling for AF rhythm, sex, age, and surrogate variables. The number of surrogate variables was determined using the R surrogate variable analysis (SVA) package; surrogate variables were recalculated for each of the genes. The SVA used unsupervised models to correct for the heterogeneity in gene expressions, such as batch effects, unknown factors, library depth, or comorbidities like AF status, history of structural heart disease, etc. We controlled the false discovery rate (FDR) of the multiple testing with the R q value package. Genes were considered significantly coexpressed if q value was <0.05 (Supplemental Excel 1).

Expression correlations were also performed, limiting analyses to the genes within each gene group, i.e. the correlation of each candidate AF risk gene with all other AF candidate genes was calculated. Genes with a q value <0.05 were considered significantly coexpressed. For each gene group, agglomerative hierarchical clustering with the compete linkage method was used, where genes start as singletons and are successively joined to form one cluster that contains all the genes.

### Weighted Gene Coexpression Network Analysis (WGCNA)

WGCNA is a method to cluster genes into distinct modules by degree of connectivity determined by commonly coexpressed (as measured by correlation) genes. To construct the required correlation matrix for WGCNA, first the conditional mean (log_2_ scale) of LAA gene expression was modeled using the limma-voom^9^ approach with an extensive set of known AF risk factors and other covariates. Residuals were calculated from this model to remove mean effects and used as inputs to the RSVP^10^ approach for estimating correlation matrices in the presence of latent factors. The RSVP adjusted correlation matrix was then normalized using the spqn approach^11^ to remove the mean-variance bias common in RNA-seq data due to differential measurement error. A weighted gene correlation network was built using the RSVP-spqn adjusted and normalized correlation matrix using the CEMiTool R package^12^. Hub genes were defined as genes in the top 1% for intra-module connectivity (Supplemental Excel 2).

### Candidate pathway identification

Canonical pathways, upstream regulators, and toxicity lists were identified using the Ingenuity Pathway Analysis (IPA) software package (QIAGEN, Inc.). IPA uses gene expression data to predict biological pathways that are activated by comparing known genes within a set of pathways to expression patterns in the imported dataset. Ensemble ID number, q-value, and log fold change values for coexpressed genes were imported into IPA. Coexpressed genes with q values <0.05 were used for analysis. Significance of association between the data and canonical pathways was determined by the ratio of the number of overlapping genes from our dataset to the total number of IPA dataset genes in a particular pathway, to determine the most significantly affected pathways. Fisher’s exact test was used to determine the p-value of association between genes and the canonical pathway, where p <0.05 was considered significant. Toxicity lists are generated based on sets of molecules that are known to be involved in a particular type of toxicity. IPA analyses were performed for the coexpression partners of each gene in the All Candidate Genes group (Supplemental Excel 3-5), and for each module from the WGCNA (Supplemental Excel 6). Over representation analysis (ORA) was performed on each WGCNA module using the CEMiTool package^12^ and the Molecular Signatures Database^13^ v7.1 (Supplemental Excel 7).

## RESULTS

### Patient characteristics of the 235 subjects have been previously described^3^

#### AF GWAS Loci SNPs

AF GWAS risk SNPs from two recently published meta-analyses were aggregated to create a comprehensive list of 166 SNPs: 142 risk variants were identified by Nielsen, et al^6^, with an additional 24 independent risk variants identified by Roselli, et al^5^. The 166 SNPs were located in 135 distinct loci (Supplemental Excel 8). The locus with the most independent risk SNPs^6^ was at 4q25, near *PITX2*, which had 10; loci labelled *SCN10A, SCN5A, KCNN3, NEURL1, RPL3L*, and *LINC01142* each had 3; loci *LRRC10, NKX2-5, ZFHX3, LINC00477, HSF2, NACA, AKAP6*, RP11_380D231 (AC098798.1), *FBXO32, MEX3C* had 2. All remaining loci were associated with 1 risk SNP.

#### AF Candidate Gene Lists (Table 1)

Of the 166 risk variant SNPs from the published meta-analyses, 80 SNPs had significant cis-eQTL association (q<0.05) with 131 genes from our LAA mRNA-Seq eQTL European-ancestry database comprising the Any eQTL Genes group. Of the 131 genes in this group, 83 genes had not been previously annotated by Nielsen, et al^6^, as potential AF genes (Table 2). The functions of many of these genes are not yet known.

**Table 1.**
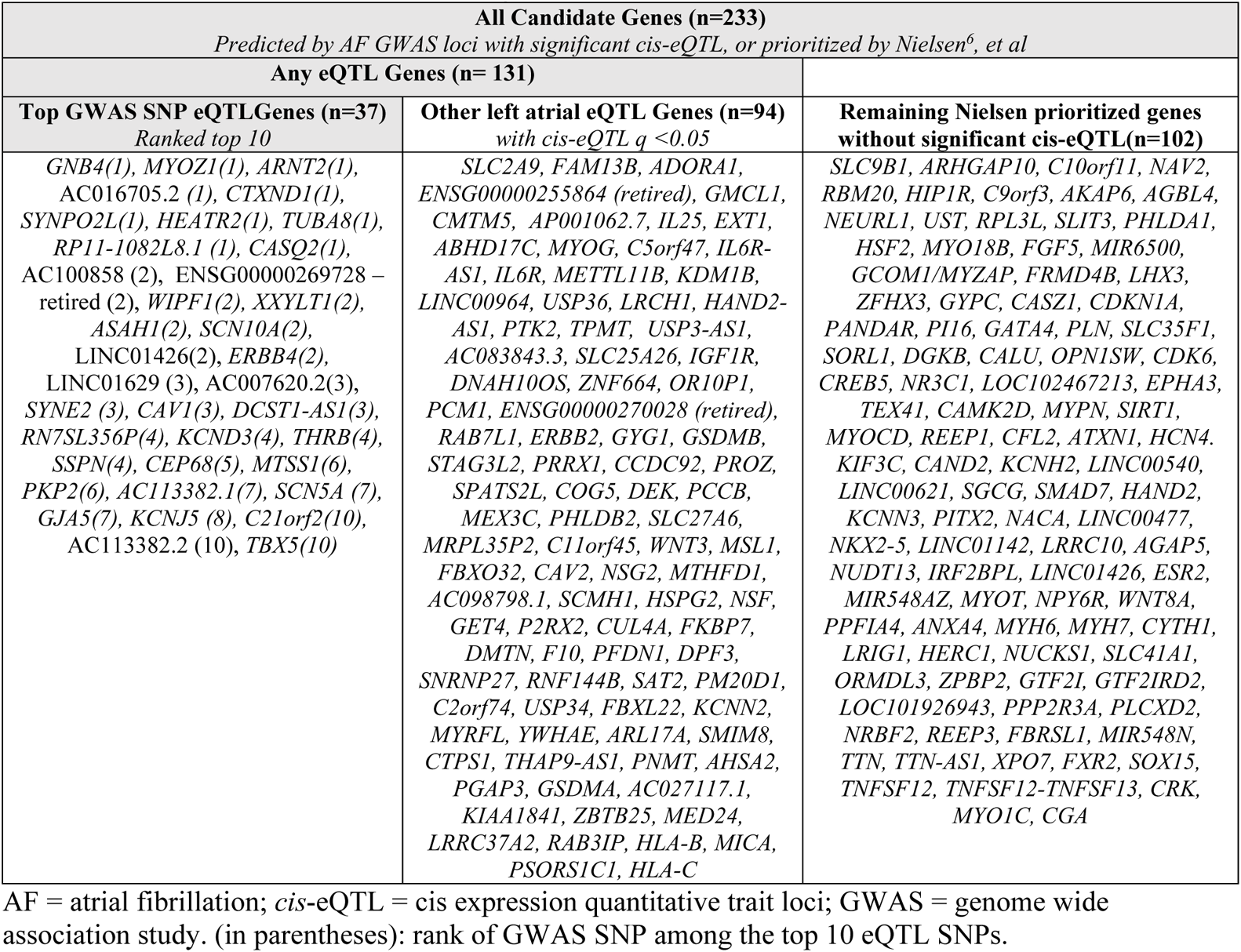
Candidate AF genes. Shaded headings indicate the 3 targeted groups of study genes. Genes are listed in order of statistical significance of eQTL SNPs.

**Table 2.**
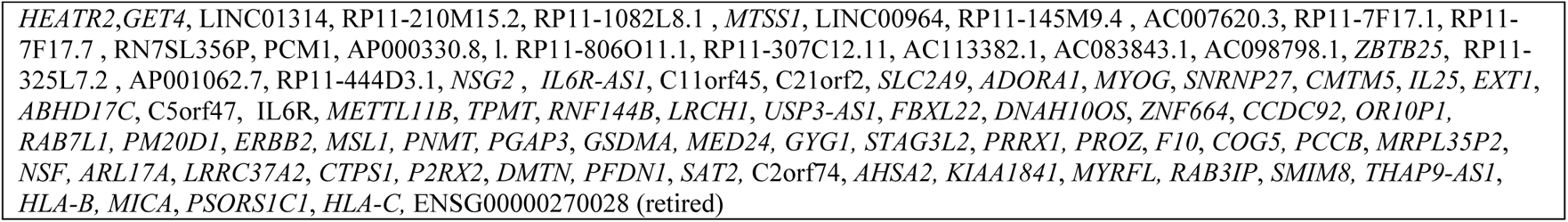
Genes identified by significant left atrial cis-eQTLs but not prioritized by Nielsen, et al ^6^.

The All Candidate Genes group comprised the 131 genes from the Any eQTL Genes group and 102 genes prioritized by Nielsen, et al^6^ (Table 1). Of the 233 genes, 111 are known protein-coding genes (Supplemental Figure 1), encoding mostly enzymes and transcription factors with predominant localization in the cytoplasm (32%) or nucleus (22%) and the remaining in the plasma membrane (17%), extracellular space (7%) or other (22%). By function (Supplemental Figure 1), 12% encode transcription regulators, 12% enzymes, 5% transporters, 4% ion channels, 4% kinases, 3% peptidases, and 2% transmembrane receptors.

#### Genes Coexpressed with Each Candidate Gene

Of 233 candidate genes in the All Candidate Genes group, 5 AF candidate genes were not significantly expressed in our human adult LAA tissues (MIR6500, *PANDAR*, MIR54810, MIR548AZ, LOC101926943) and 3 (ENSG00000269728, ENSG00000255864, ENSG00000270028) were retired by Ensembl.

Coexpression partners of the remaining 228 AF candidate genes ranged from 0 to 1928 genes. C4orf32 had the highest number of coexpressed genes (1928 genes), followed by *FXR2*, which was significantly coexpressed with 1889 genes.

#### Novel candidate genes identified by coexpression analyses

Adenosine monophosphate (AMP) activated protein kinase (AMPK) alpha 2 catalytic subunit (*PRKAA2*) and calcium/calmodulin-dependent protein kinase II beta (*CAMK2B*) were the most common genes co-expressed with the AF candidate genes that are not themselves GWAS-identified. Other commonly coexpressed genes included prostaglandin F2 receptor inhibitor (*PTGFRN)*, talin 1 (*TLN1)*, sarcolemma associated protein (*SLMAP)*, PDZ and LIM domain 5 (*PDLIM5)*, and methionine sulfoxide reductase B3 (*MSRB3)* (Supplemental Figure 2).

#### Top Canonical Pathways of Coexpressed Genes

IPA was used to identify top candidate pathways of each group of genome-wide coexpressed partners for each gene in the All Candidate Genes group (Supplemental Excel Folder10). After ranking IPA canonical pathways for each candidate gene by p-value, the top 5 significant canonical pathways were listed for each gene (Supplemental Excel 9). Mitochondrial dysfunction, oxidative phosphorylation, epithelial adherens junction signaling, sirtuin signaling pathway and actin cytoskeleton pathway were the most frequent pathways listed among the top 5 canonical pathways (Supplemental Excel 3). For the Any eQTL Genes group (Supplemental Excel 4), epithelial adherens junction signaling, mitochondrial dysfunction and oxidative phosphorylation, germ cell-Sertoli cell junction signaling and axonal guidance signaling were the most frequent pathways listed. For the Top GWAS SNP eQTL Genes group (Supplemental Excel 5), EIF2 signaling, axonal guidance signaling, sertoli cell-sertoli cell junction signaling, regulation of eIF4 and p70S6K signaling, oxidative phosphorylation, and mitochondrial dysfunction were the most frequent pathways listed among the top 5 canonical pathways. Overall, the most frequent pathways were those involved in energy and metabolism signaling, including mitochondrial dysfunction, oxidative phosphorylation, and sirtuin signaling. Stress and injury signaling pathways were also represented and included EIF2 signaling pathways, regulation of eIF4 and p70S6K signaling, protein ubiquitination and cardiac hypertrophy signaling.

Top canonical pathways, top upstream regulators and top networks from pathway analyses within each of the gene groups are shown in Supplemental Table 1.

#### WGCNA Modules

WGCNA characterized 64 distinct modules from 17,357 genes expressed in the LAA (Supplemental Excel 2). 48 of these modules, including a module containing uncorrelated genes (‘not_correlated’), contained at least one AF candidate gene. Candidate AF genes were overrepresented in 8 modules (p<0.05; modules 21, 2, not correlated, 25, 11, 42, 39, 24). Hierarchical clustering (Figure 1) of the modules’ top eigengene produced 15 clusters. A list of genes for each module, alongside with hub genes that were determined by module connectivity for each module are available in Supplemental Excel file 2.

**Figure 1.**
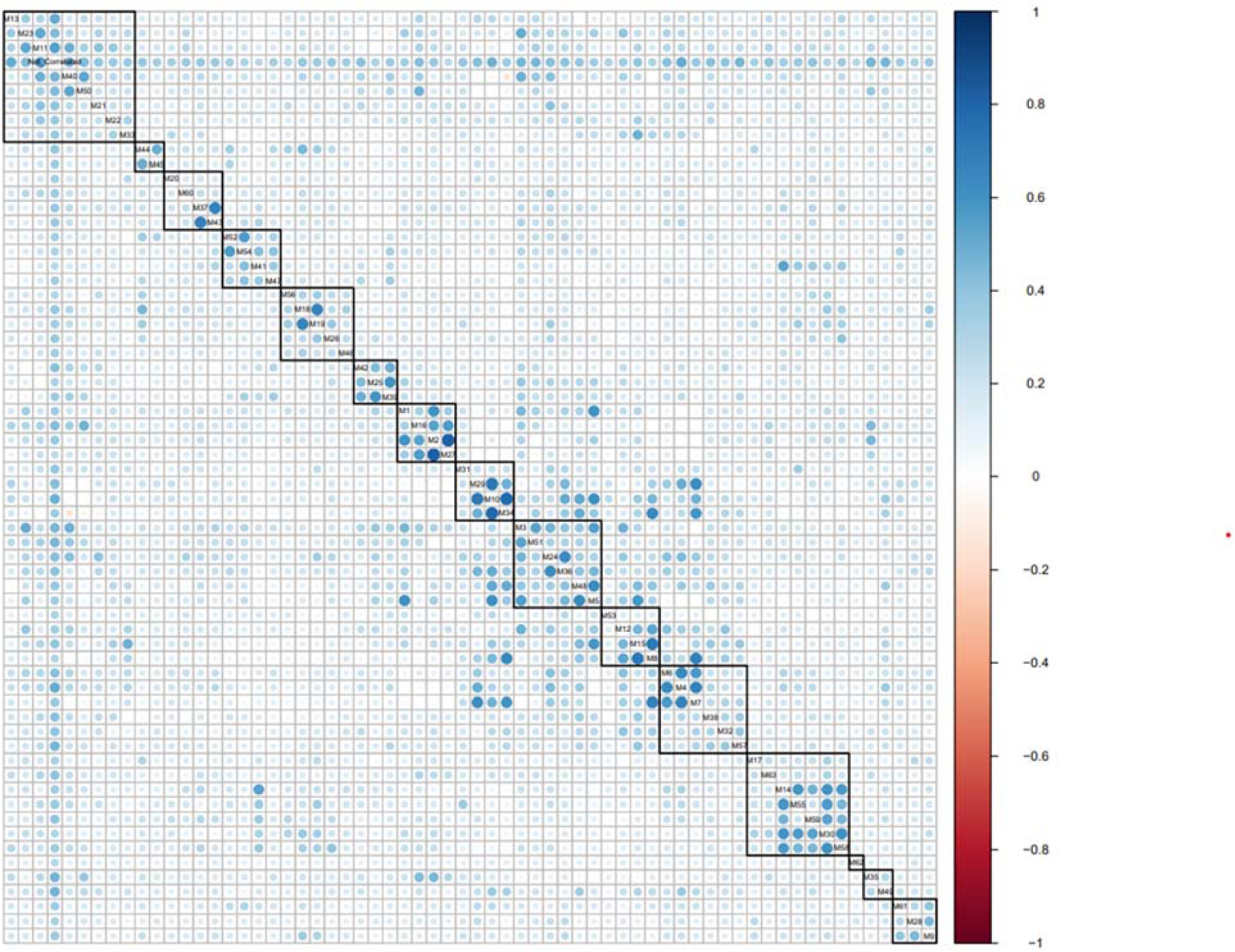
WGCNA correlation plot. WGCNA identified 64 distinct modules, which included a ‘non correlated’ module. 8 of the 48 modules had over-representation of AF genes in their respective modules. Hierarchical clustering of the modules show clustering of AF overrepresented modules 25, 39 and 42, and clustering of modules 11, 21, and ‘not correlated’. The size and shade of circle reflect strength of correlation, and color shows direction of correlation.

Concordant with IPA analyses, the modules with AF risk gene overrepresentation suggest that AF genes are over represented in modules related to intracellular ion homeostasis, metabolic/energy utilization, gene expression regulation, and signaling pathways of stress and unfolded protein responses.

The clustering of modules 25, 39, and 42 (Figure 1), suggests that genes in these three modules are closely related. Gene set enrichment analyses of these 3 modules (Supplemental Excels 6 & 7) suggest that genes in these modules are involved in remodeling of the heart through a combination of myogenesis and the unfolded protein response.

#### Biological pathways of top AF candidate genes

Coexpression analyses can provide insight into biological function of top AF candidate genes. These include transcription factors such as *PITX2*, *ZFHX3*, top implicated genes by AF GWAS for which function remains elusive, as well as candidate causal genes in the Top GWAS SNP eQTL Genes group. IPA pathways for *PITX2* (Figure 2) and *ZFHX3* (Figure 3) are shown.

**Figure 2.**
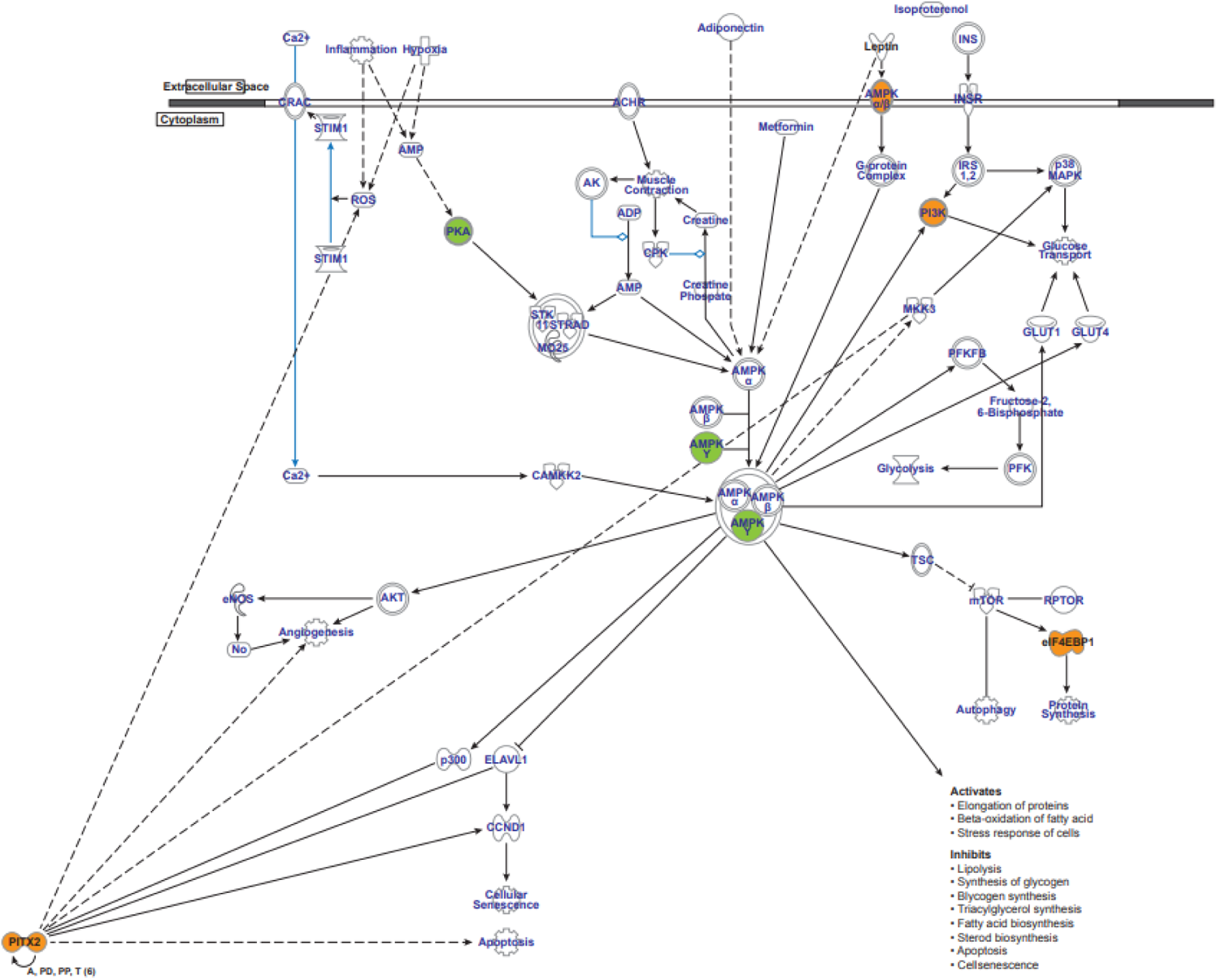
PITX2 and coexpressed genes signaling pathways. Top canonical pathways include protein handling (e.g. β-alanine degradation, valine, 4-aminobutyrate and glutamine degradation) and and major signaling pathways (e.g. PI3K/AKT, G protein coupled receptor signaling, IGF-1, PDGF signaling, renin-angiotensin and AMPK signaling pathways). PIK3 is critical in insulin resistance and in the regulation of myogenic differentiation, myoblast proliferation, and myotube hypertrophy.Activationof the PI3K-Akt signaling pathway, mediated through IGF-1, decreases autophagy and apoptosis. AMPK activity and the resultant glycogen accumulation is correlated with changes in metabolic pathways and Akt phosphorylation.

**Figure 3.**
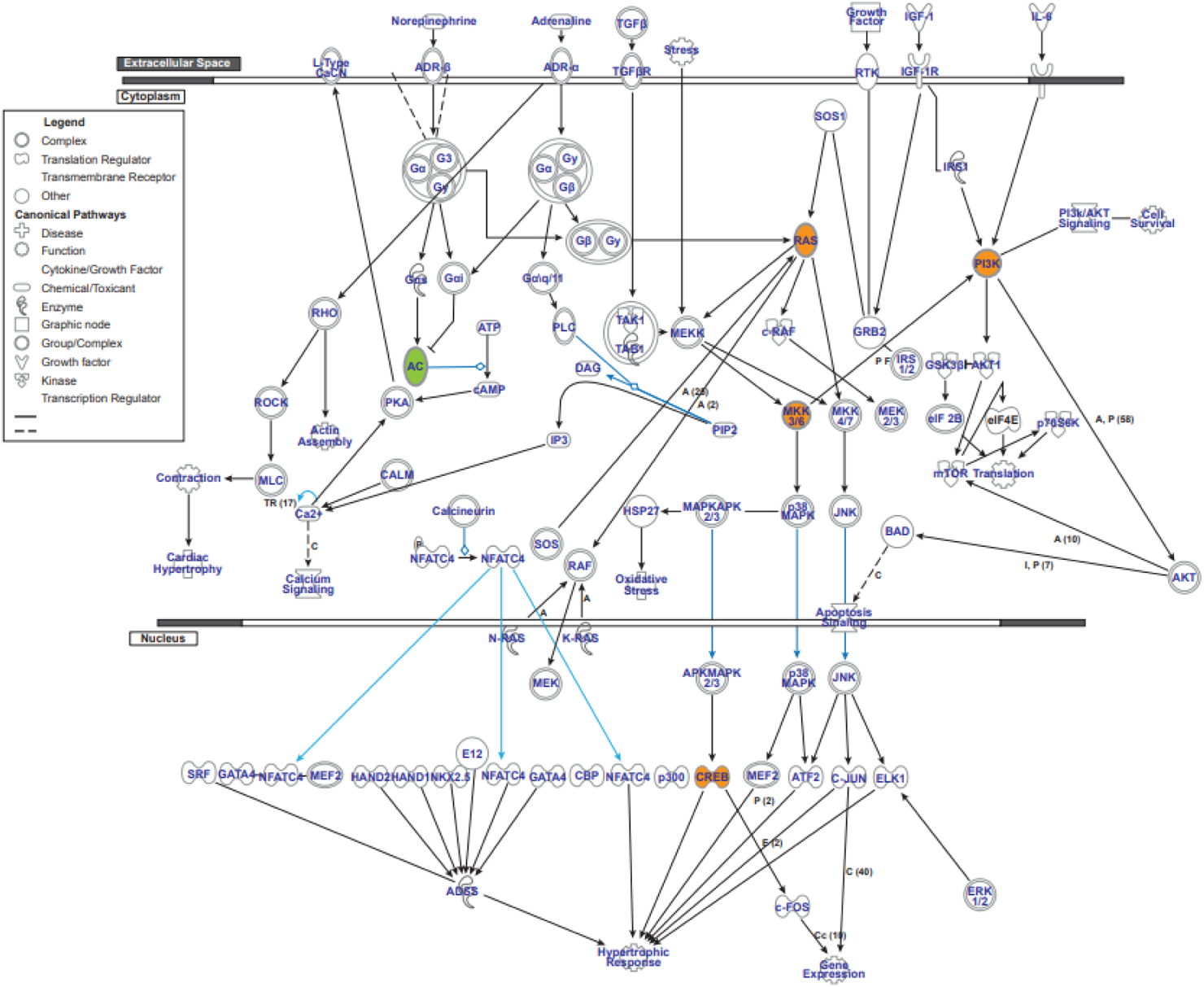
ZHFX3 and coexpressed genes signaling pathways. *RAS*, *PI3K* and *CREB* have been linked to modulation of cardiomyocyte apoptosis.

## DISCUSSION

Despite having identified over 135 genetic loci associated with AF from prior meta-analyses, establishing functional connections and causal genes contributing to AF pathophysiology remains challenging. Here we utilized human atrial eQTL analyses to identify top candidate causal AF genes. To advance our understanding of the biological mechanisms of these candidate genes, we identified gene coexpression partners for each candidate gene and WGCNA modules that inform functional characterization using pathway and gene set analyses. This application provides insight into the function of genes nearest the top AF GWAS loci, for example *PITX2* and *ZFHX3*, for which connections to AF mechanisms remain challenging.

### PITX2

The top locus implicated by AF risk GWAS is on chromosome 4q25. The nearest gene is *PITX2*^5, 6^, which encodes a transcription factor that regulates left-right asymmetry during organ morphogenesis, suppresses a sinus node program in the LA, and is critical to formation of the pulmonary veins^14^, making it an attractive candidate causal gene. *PITX2* is active during cardiac development and decreases with maturation, likely contributing to our failure to identify significant eQTLs in our adult LAA tissues. Nevertheless, *PITX2* remains expressed in our adult LA tissues, though in lower levels in patients with AF,^3^ suggesting an additional function(s) in the adult LA. Top canonical pathways of genes coexpressed with *PITX2* (Figure 2) include amino acid metabolism (e.g. alanine degradation, valine, 4-aminobutyrate and glutamine degradation, driven by *ABAT* and *ALDH6A1*), and major signaling pathways, such as PI3K/AKT, G protein coupled receptor signaling, IGF-1, PDGF signaling, renin-angiotensin and AMPK signaling pathways, all commonly mediated by the phosphoinositide-3-kinase regulatory subunit 1 (*PIK3R1*), and also AMPK non-catalytic subunit γ 2 (*PRKAG2)*. *PIK3R1* encodes for a subunit of PIK3, which is critical in insulin resistance^15^, and in the regulation of myogenic differentiation, myoblast proliferation, and myotube hypertrophy^16, 17^. The PI3K-Akt signaling pathway is activated by insulin like growth factor-1 (IGF-1)^17^ which is critical in regulation of energy metabolism in cardiomyocytes^18^, and decreases autophagy and apoptosis^19–22^. Inhibition of the PI3K-Akt signaling in the heart is thought to contribute to AF in patients on ibrutinib^23^, an immunotherapeutic agent that acts as a Bruton’s tyrosine kinase inhibitor ^24^. PRKAG2 is one of three AMPK regulatory subunits, and is abundant in the heart^25^. *PRKAG2* gain of function mutations increase AMPK activity and promote accumulation of glycogen in cardiac tissues via transcriptional changes in glucose handling regulators^26^. *PRKAG2* mutations are considered a metabolic storage disease and not a disease of the sarcomeres - this is further evidenced by the absence of fibrosis or myocyte disarray in patients with a mutation presenting with either hypertrophic cardiomyopathy and/or conduction abnormalities^27^. Increased AMPK activity and the resultant glycogen accumulation are also correlated with changes in metabolic pathways and AKT phosphorylation^26^. The AKT signaling pathway also regulates myocyte hypertrophy^28^. These findings suggest that *PITX2* may play a role in metabolic energy utilization and/or regulation in the heart, and dysregulation may cause metabolic dysregulation and imbalance in autophagy, with subsequent disease.

IPA analysis also highlighted fibrotic signaling pathways, driven by transmembrane protein 67 (*TMEM67)*, cannabinoid receptor 1 *(CNR1)*, collagen type IV α2 chain *(COL4A2)*, insulin like growth factor binding protein 5 *(IGFBP5)*, adrenergic receptor β2 (*ADRB2*), plasminogen activator, urokinase *(PLAU)*, angiotensin I converting enzyme *(ACE)*, neurofibromin 1*(NF)*, and Xin actin binding repeat containing 1 *(XIRP1)*. AMPK activation inhibits Transforming Growth Factor (TGF) beta signaling and target transcripts of pathways associated with cardiac fibrosis^26^. Although *PITX2* had no prior known role in fibrosis signaling, this network pattern suggests that *PITX2* may suppress general fibrosis and tissue remodeling pathways, perhaps by the AMPK pathway; *PITX2* loss of function may promote activation of fibrosis pathways, which may predispose to AF.

*PITX2* is most strongly coexpressed with *PITX2* adjacent non-coding RNA (*PANCR*), which we reported is an upstream regulator of *PITX2*^29^. Both are located in module 58, which contains genes encoding ion channels (*SCN4B, ANO1, KCNJ8, CACNA1H, P2RX3, KCNT1, SCN3A*), cytoskeletal proteins (*MYH9, ADCY3, MYO1B, MYO6*), and G protein coupled receptors. These associations suggest further functional impact of *PITX2* on these pathways in adult atria.

### ZFHX3

*ZFHX3* is associated with the regulation of numerous pathways, including growth and differentiation of neuronal tissue^30^, regulation of inflammation^31^, and tumor suppression through regulation of *MYC* transcription^32^. *ZFHX3* has been implicated in calcium homeostasis, and knock down of *ZFHX3* promotes arrhythmogenesis in atrial myocytes,^33^ mediated by increased expression of sarcoplasmic/endoplasmic reticulum Ca2+ATPase 2a (SERCA2a), ryanodine receptors, ultra-rapid delayed rectifier potassium currents, transient outward currents and acetylcholine-sensitive potassium currents.

WGCNA analysis associates *ZFHX3* with module 2, which was overrepresented by AF risk genes. Hub genes of module 2 include rho guanine nucleotide exchange factor 12 (*ARHGEF12*), phosphoinositide kinase, FYVE-type zinc finger containing (*PIKFYVE*), and kinesin family member 1B (*KIF1B*). Gene enrichment analyses suggest module 2 likely contains genes critical in mitochondrial function modulation and apoptosis.

The top canonical pathways of *ZFHX3* and coexpressed genes include cell proliferation pathways such as acute myeloid leukemia, and prostate cancer signaling pathways, driven by *KRAS*, mitogen-activated protein kinase kinase 6 (*MAP2K6*), phosphatidylinositol-4,5-bisphosphate 3-kinase catalytic subunit α (*PIK3CA*), cAMP responsive element binding protein 1 (*CREB1*), and phosphatase and tensin homolog (*PTEN*). *RAS*, *PI3K* and *CREB* gene expression is positively associated with *ZFHX3* (Figure 3). As these genes have been linked to modulation of cardiomyocyte apoptosis,^19, 22, 34^ *ZFHX3* may also regulate apoptosis and hypertrophy of cardiomyocytes. This is consistent with the top toxicity lists suggested by IPA which include cardiac necrosis/cell death, cardiac fibrosis and hypertrophy, and followed by RAR activation, and NRF2-mediated oxidative stress.

### Identification of Novel AF Candidate Genes by Coexpression Analyses

Identification of genes coexpressed with GWAS-identified AF risk genes provides novel insights into AF. While not identified by GWAS as AF risk genes, these genes may also play significant roles in AF pathogenesis or yield additional targets for functional studies. Based on frequency of being coexpressed with the putative AF genes, we identified two genes of potential interest in AF pathophysiology: *<u>PRKAA2</u>*, the α2 catalytic subunit of AMPK, and *<u>CAMK2B</u>*, a subunit of calcium/calmodulin-dependent protein kinase II (*CAMKII*).

#### PRKAA2

is a catalytic subunit of AMPK in the heart^35^. AMPK plays an important role in cellular energy and metabolism sensing and regulation. Activation of AMPK induces mitochondrial biogenesis, as well as increases insulin sensitivity and fatty acid oxidation^36^. AMPK activity is induced by drugs, such as metformin and thiazolidinediones^37^. Metformin affects phosphorylation of PRKAA2^37^. Activity of PRKAA2 increases with muscle contraction and exercise^35^. *PRKAA2* deletion causes defects in autophagy, mitochondrial fission and fragmentation, and insulin resistance^38^ in mice. In the heart, PRKAA2 deletion increases release of atrial natriuretic peptide, myocardial fibrosis and hypertrophy, with subsequent decrease in ejection fraction^39^ and arrhythmogenesis^40^. In a study of 645,710 diabetics, metformin was associated with 20% less AF^41^. *PRKAA2* is located in module 1. GSEA and IPA analyses suggest genes in module 1 may be involved in cell death signaling.

#### CAMK2B

is a subunit of calcium/calmodulin-dependent protein kinase II (*CAMKII*) that is expressed primarily in the brain, followed by the heart. In the heart *CAMKII* is activated by β-adrenergic signaling, Gq coupled receptor signaling, presence of reactive oxygen species, hyperglycemia, ischemic injury, and other stimuli that cause increased intracellular calcium^42^.

Persistent activation of *CAMKII* promotes arrhythmogenesis, myocardial apoptosis and necroptosis^43^. In contrast to apoptosis that requires ATP derived from mitochondria, necroptosis is mediated by opening of the mitochondrial permeability transition pore (mPTP), resulting in dissipation of mitochondrial membrane potential and subsequent shutdown of the mitochondria^42^. Necroptosis triggers an inflammatory response that results in tissue injury and likely contributes to many cardiovascular disease processes ^42^. CAMK2B is located in module 22 which is associated with genes involved in myogenesis and development.

### WGCNA modules with overrepresentation of AF risk genes

AF genes are over represented in modules related to intracellular ion homeostasis, metabolic/energy utilization, gene expression regulation, and signaling pathways of stress and unfolded protein response (Table 3).

**Table 3.**
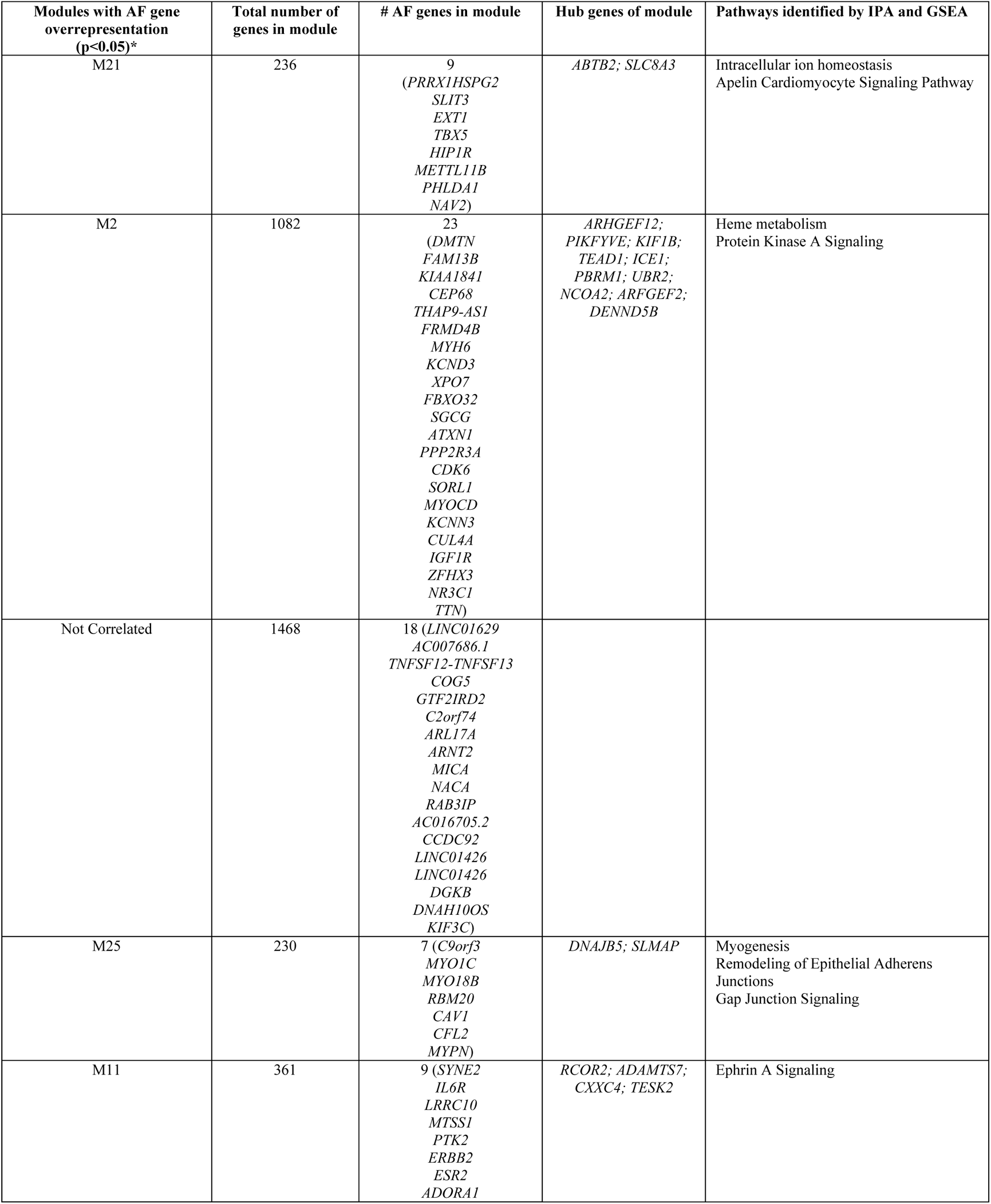
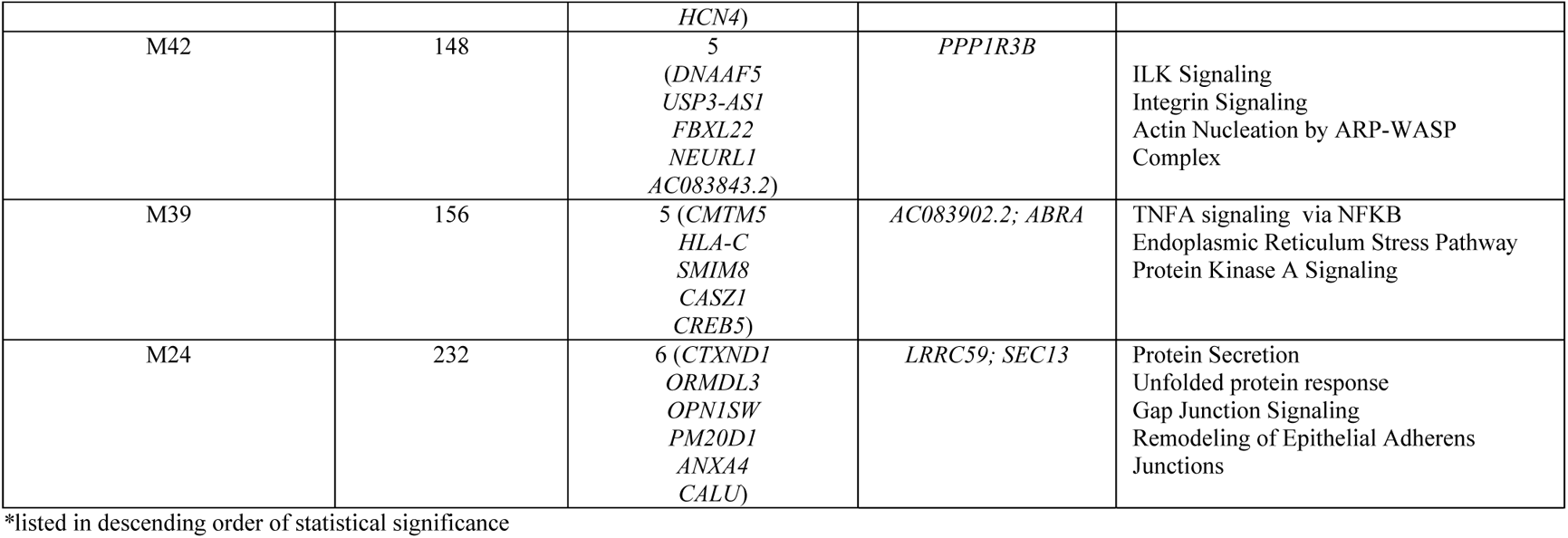

One of the most statistically significant modules with AF over representation is module 2. Heme metabolism is the hallmark pathway from GSEA for module 2. Iron deficiency is seen in patients with heart failure, and portends higher mortality^44^. In contrast, iron overload can result in AF^45^. Iron homeostasis affects mitochondrial function and ATP-linked respiration by modulating activity of electron train complexes I, II and III^46^. Respirometry studies from human LAA tissues of patients with AF and diabetes undergoing coronary artery bypass graft surgery also implicate impairment of Complexes I and II^47^. In rapidly paced cardiomyocytes, mitochondrial dysfunction has been linked to AF progression, mediated by activation of autophagy and endoplasmic reticulum stress induction^48^. One of the top canonical pathways from IPA analysis of module 1 includes Protein Kinase A (PKA) signaling. Oxidative stress activates PKA signaling in cardiac myocytes which triggers apoptosis and reduced mitochondrial respiration^49^. PKA activation indirectly inhibits mitochondrial fission and increases mitochondrial fusion, with resultant superoxide production and altered mitochondrial potential with increase in mitochondrial pore permeability^50^. Analysis results of module 2 from both IPA and GSEA suggest a potential significant role of the mitochondria in AF.

These examples demonstrate how WGCNA modules and pathway analyses of coexpressed genes can help elucidate potential function of AF risk genes, including those for which mechanistic connections to the disease have remained poorly defined. Our overall findings are consistent with the two meta-analyses reported by Roselli et al^5^, and Nielsen et al.,^6^ but are also distinct from previous findings. Roselli et al^5^ noted themes of cardiac development, electrophysiological, and cardiomyocyte contractile or structural functional groups. Nielsen et al.^6^ noted cardiac structural remodeling as a recurrent theme and postulated that genes active during fetal heart development are reactivated in response to stress in the adult heart. We also see themes related to cardiac structural change, but also discovered involvement of unfolded protein response, mitochondrial and metabolic pathways, especially those of AMPK.

### Dual Risk Model for Development of AF

Differences between the All Candidate Genes group and the Any eQTL Genes group highlight the differing expression patterns that may be present in cardiac development compared to later in life, when AF typically presents. The All Candidate Genes group contains GWAS implicated genes active in development. The Any eQTL Genes group indicate genes that are active in adult LAA, whose expression may serve as a barometer of function in the adult atria. These findings support our proposed dual risk model for development of AF (Figure 4)^51^. In this model, genes active during cardiac structural development contribute to genetic susceptibility to AF, for example potentially by alterations in pulmonary vein development (e.g. *PITX2*) that may contribute to pulmonary vein triggers. However, with exposure to stresses later in life, transcriptional responses to cellular stress, such as metabolic stress, mitochondrial dysfunction, oxidative stress, sirtuin signaling and abnormal proteostasis may lead to altered, overwhelmed or reduced transcriptional responses, as suggested by our prior study of differential gene expression in AF^52^ and the current analyses. Indeed, AF is associated with heterogeneous cardiovascular stressors, including aging, obesity, hypertension, coronary or valvular disease, heart failure, metabolic syndrome, sleep apnea, excessive alcohol, and extreme exertion. Thus, in genetically susceptible individuals, the imbalance in transcriptional responses to stress may predispose to AF occurring later in life. In addition, the imbalance or overcompensation during response to injury, initially adaptive, may eventually lead to fibrosis and adverse remodeling, with eventual presentation of the disease. This concept may explain why AF tends to occur later in life as evidenced by the top canonical pathways in this study, including pathways involved in mitochondrial dysfunction, oxidative phosphorylation and sirtuin signaling. Metabolic stressors and pathways likely play a strong role in AF and suggest that AF occurrence may reflect crossing the threshold of cellular resilience and adaptation.

**Figure 4.**
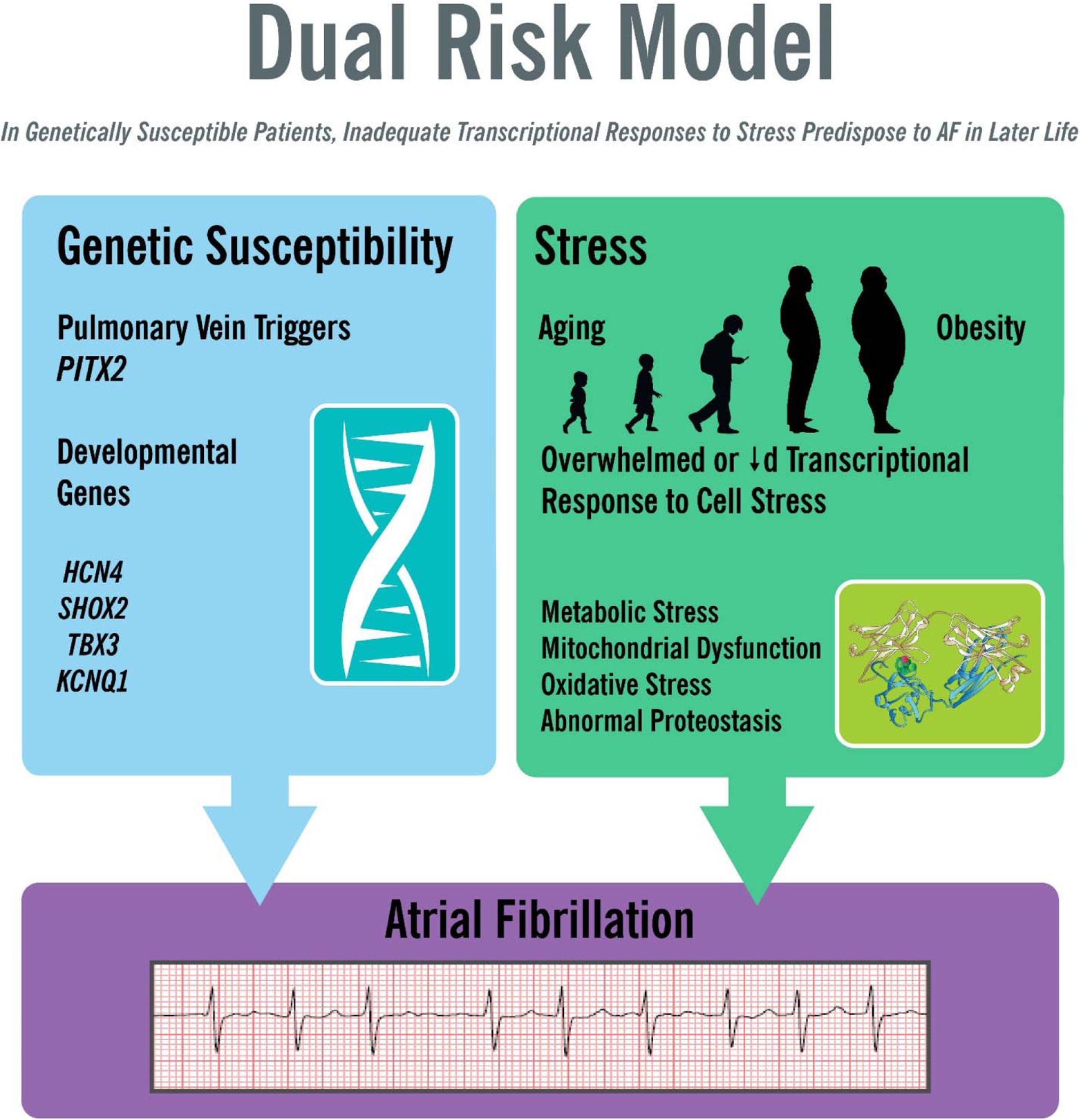
Dual Risk Model for AF. Genes active during cardiac development may increase genetic susceptibility to AF. Stressors, such as aging and obesity, may lead to increased cellular stress, including metabolic, mitochondrial, oxidative, and unfolded protein stress. The combination of genetic developmental susceptibility and accumulated stressors may lead to overwhelmed or inadequate transcriptional responses which predisposes to AF occurrence.

### Limitations and Future Studies

IPA was initially designed for use in cancer data sets and relies upon curated pathways, thus our results may be skewed by the composition of the pathways. Some of these pathways may not accurately reflect the pathology of cardiovascular diseases. Nevertheless, many of the top canonical pathways seem relevant to AF. Pathway analyses may oversimplify mechanisms. Functional studies, including overexpression and knockdown models, are required to establish causation. However, these analyses are intended to help guide such studies toward relevant candidate genes and pathways. Future studies are planned or underway for several of the targets identified. Our samples were from the LAA, rather than the PVs. However, PV tissue is difficult to obtain, and the LAA is directly adjacent to the left PVs and reflects atrial function in the human adult. The left atria obtained for this study were from patients that required open heart surgery, which suggests significant disease comorbidity. Here, we attempted to adjust for heterogeneity of comorbidities with use of surrogate variable adjustments.

## Conclusions

In conjunction with recent GWAS meta-analyses demonstrating 135 loci associated with risk for AF, our genetic-genomic analyses in human adult LA tissue have identified 233 candidate genes, of which 131 had significant eQTLs. Coexpression analyses of these gene sets identified stress pathways that are implicated in AF risk. These include mitochondrial dysfunction, oxidative stress, sirtuin signaling, EIF2 signaling, and protein ubiquitination pathways. WGCNA modules, and specifically those with AF candidate genes over representation, suggest remodeling in response to injury may be occurring through changes in metabolomics, myogenesis and apoptosis. We propose a dual risk model for AF, where genes active during cardiac development contribute to genetic susceptibility to AF, for example by alterations in PV development that leads to PV triggers. In genetically susceptible individuals, development of stresses later in life, such as those associated with aging or obesity, may lead to inadequate or overwhelmed transcriptional responses to stress that can predispose to AF. Here we also identify multiple genes not previously linked to AF. We have created a resource to help scientists elucidate possible functions of genes through identification of coexpressed genes with each candidate AF gene, and associated modules in the LAA via WGCNA. The components allow for better elucidation of likely AF mechanisms and potential targets for upstream therapies.

## Data Availability

Gene read counts are available in the Gene Expression Omnibus database (GSE69890)

## SUPPLEMENTAL MATERIALS

**Supplemental Figure 1.** All Candidate genes by function and location

**Supplemental Figure 2.** Most commonly coexpressed non-GWAS genes

**Supplemental Figure 3.** Top significant IPA® canonical pathways for the coexpression networks of each candidate AF gene group

**Supplemental Table 1.** Top pathways and upstream regulators within each AF candidate gene group

**Supplemental Excel 1.** Coexpression partners of 225 candidate AF genes.

**Supplemental Excel 2.** WGCNA modules-LA genes and hub genes

**Supplemental Excel 3.** All significant top canonical pathways for the coexpression networks of the All Candidate Genes (n=233 genes)

**Supplemental Excel 4.** All significant top canonical pathways for the coexpression networks of the Any eQTL Genes (n=131 genes).

**Supplemental Excel 5.** All significant top canonical pathways for the coexpression networks of the Top GWAS SNP eQTL Genes (n=37 genes).

**Supplemental Excel 6.** IPA analysis results of WGCNA modules

**Supplemental Excel 7.** GSEA of WGCNA Modules

**Supplemental Excel 8.** eQTL GWAS SNPs

**Supplemental Excel 9.** Top 5 significant canonical pathways for each candidate AF gene and coexpression partners.

**Supplemental Pdfs** IPA analysis results of candidate AF gene and coexpression partners

---

**Supplemental Figure 1.**
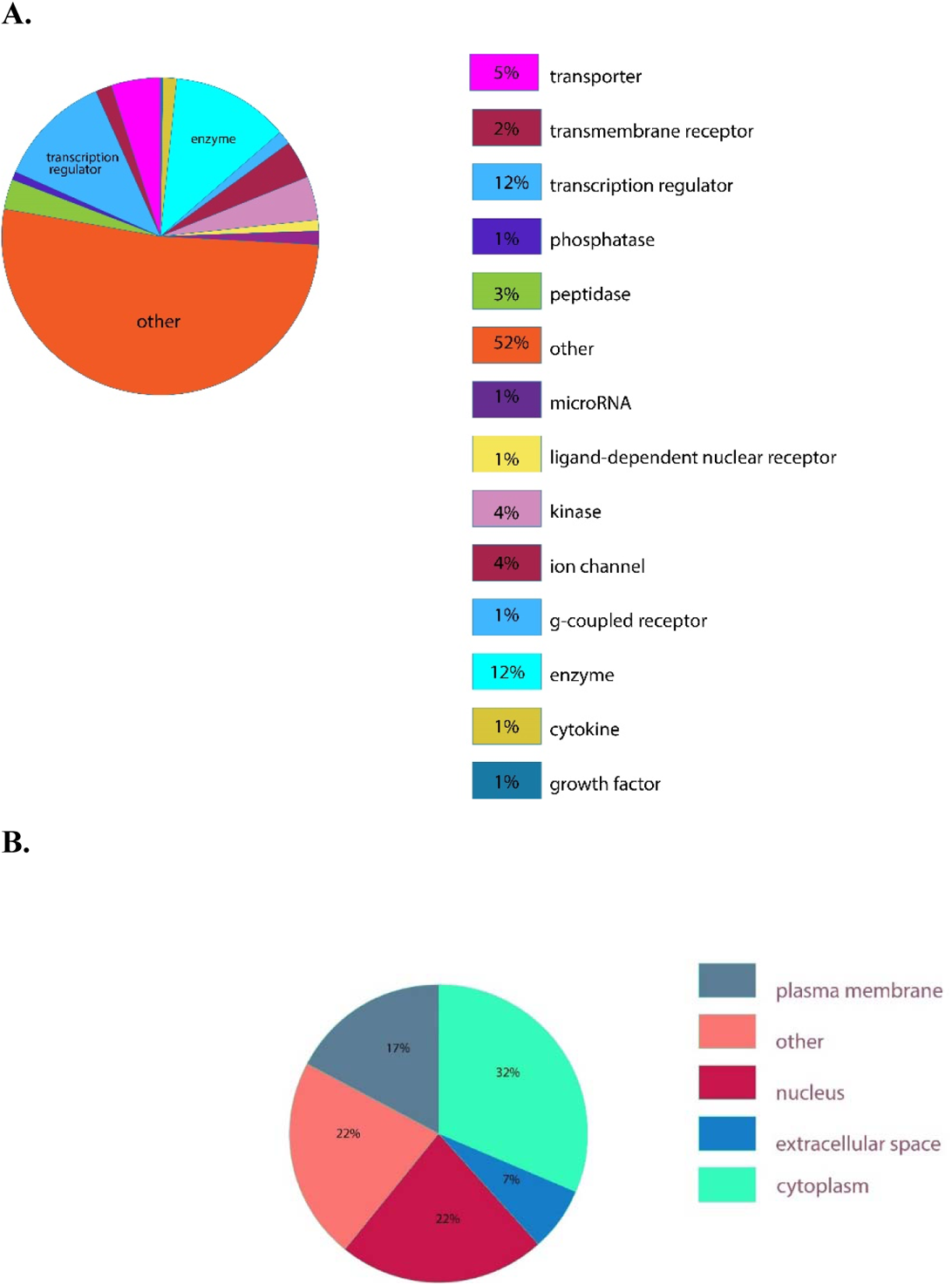
All Candidate genes by function and location. **A. Categories of Gene Function**. Of the 233 candidate atrial fibrillation genes 12% encode transcription regulators, 12% enzymes, 5% transporters, 4% ion channels, 4% kinases and 3% peptidases. **B. Categories of Gene Location.** Of the 233 candidate atrial fibrillation genes 32% are in the plasma membrane, 22% in the nucleus, 17% in the cytoplasm and 7% in the extracellular space.

**Supplemental Figure 2.**
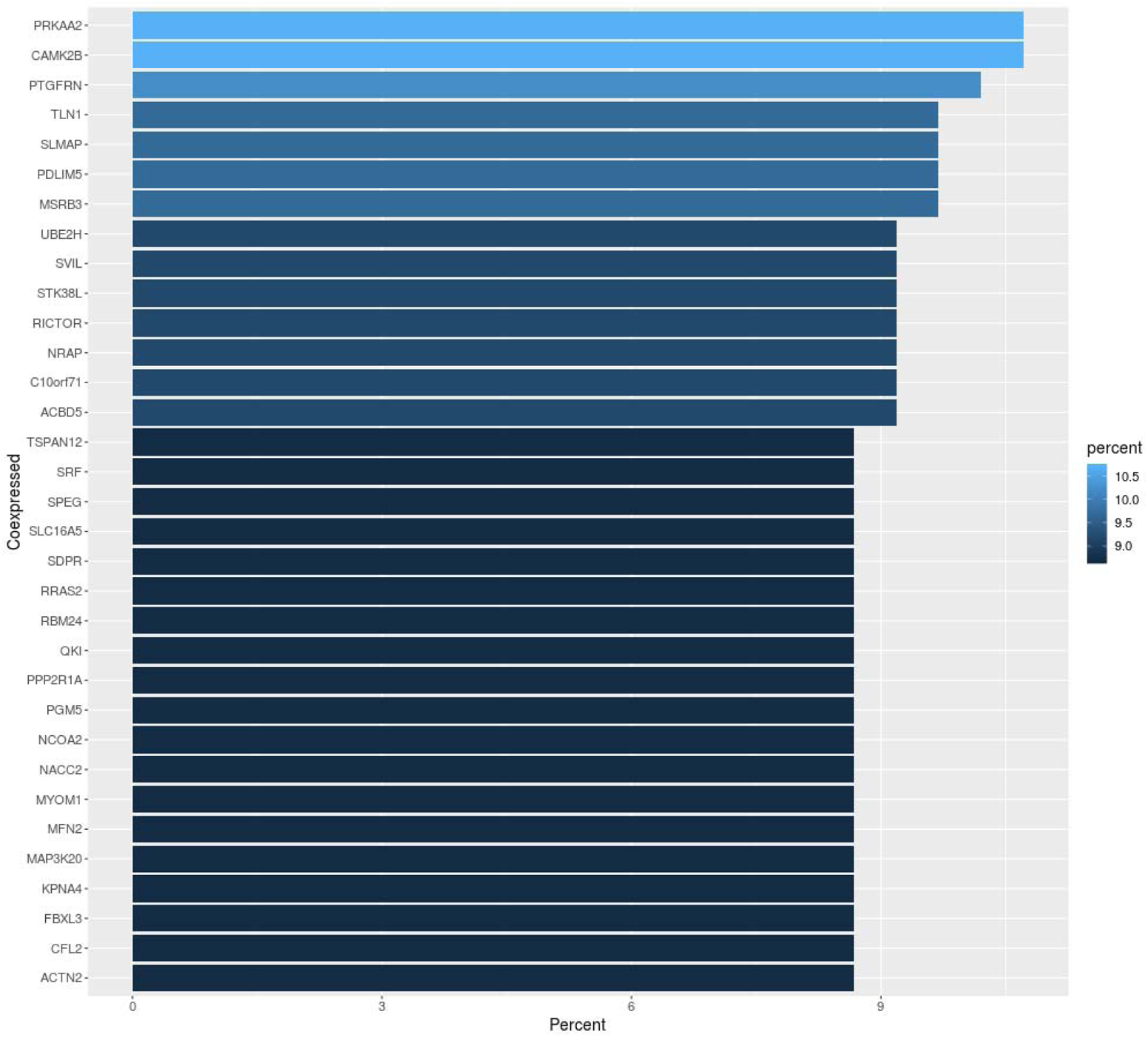
Most commonly coexpressed non-GWAS genes. *PRKAA2* and *CAMK2B* are non AF GWAS genes that are most frequently coexpressed with AF GWAS candidate genes, followed by *PTGFRN, TLN1, SLMAP*, *PDLIM5* and *MSRB3*.

**Supplemental Figure 3.**
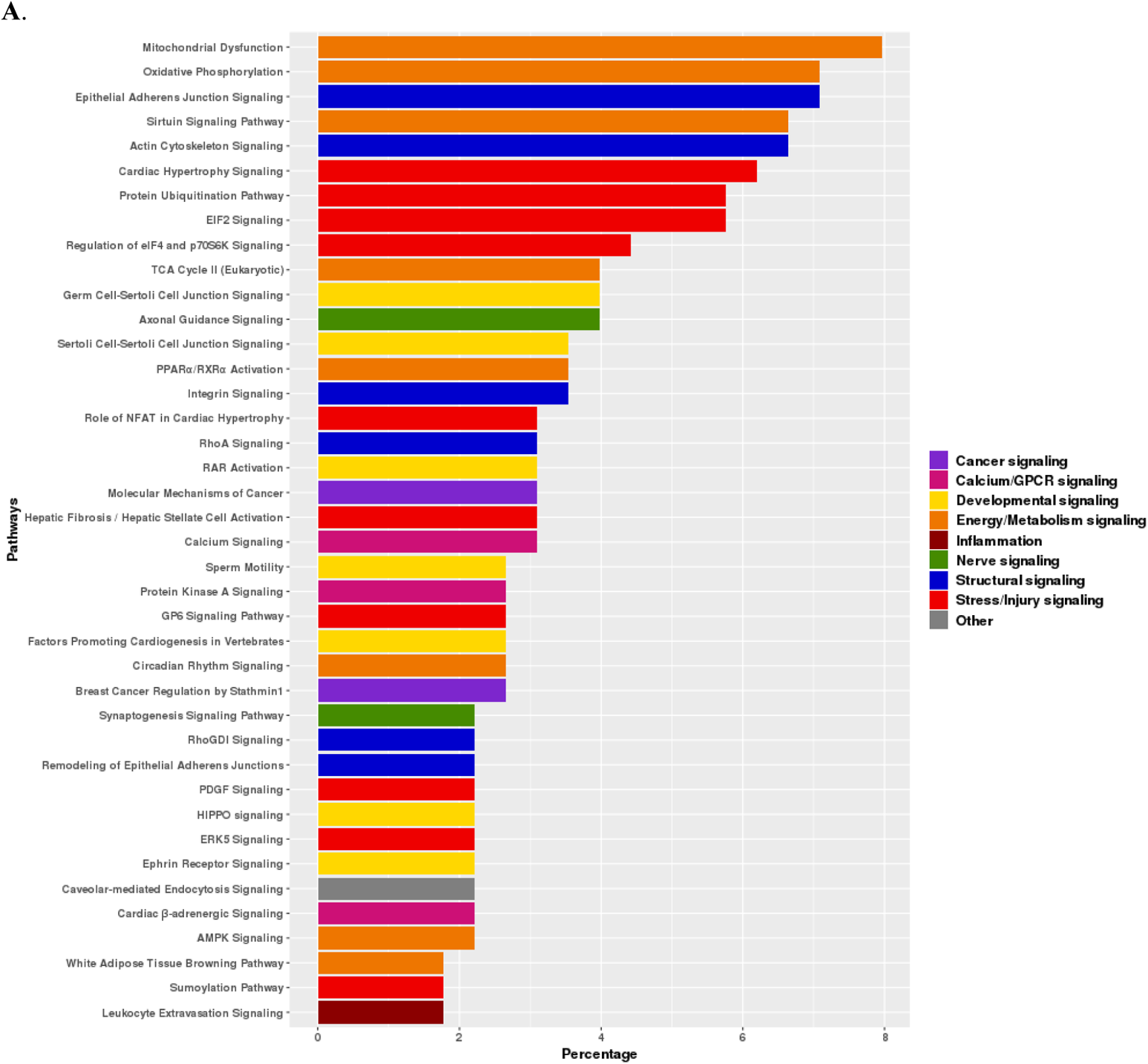
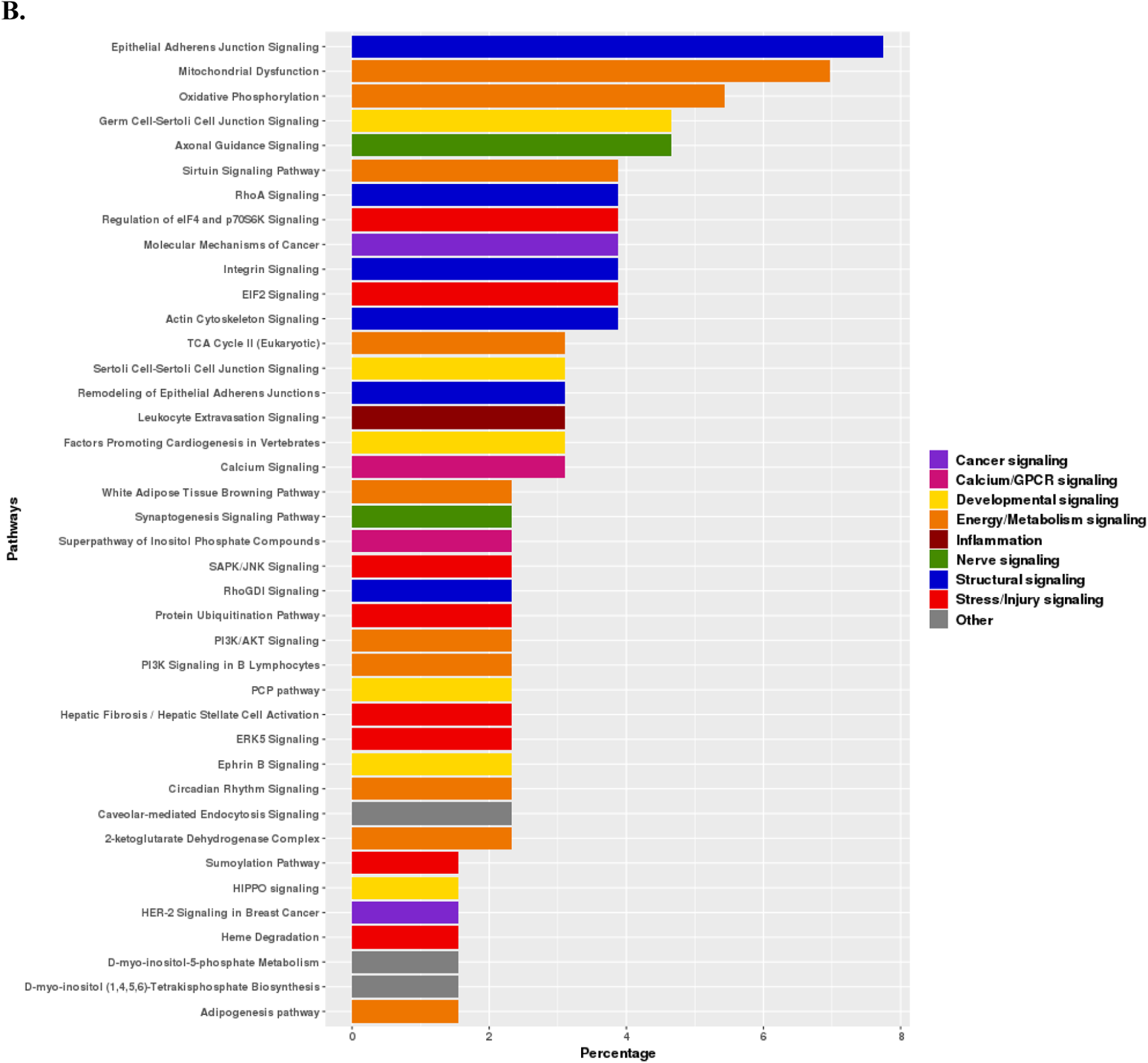
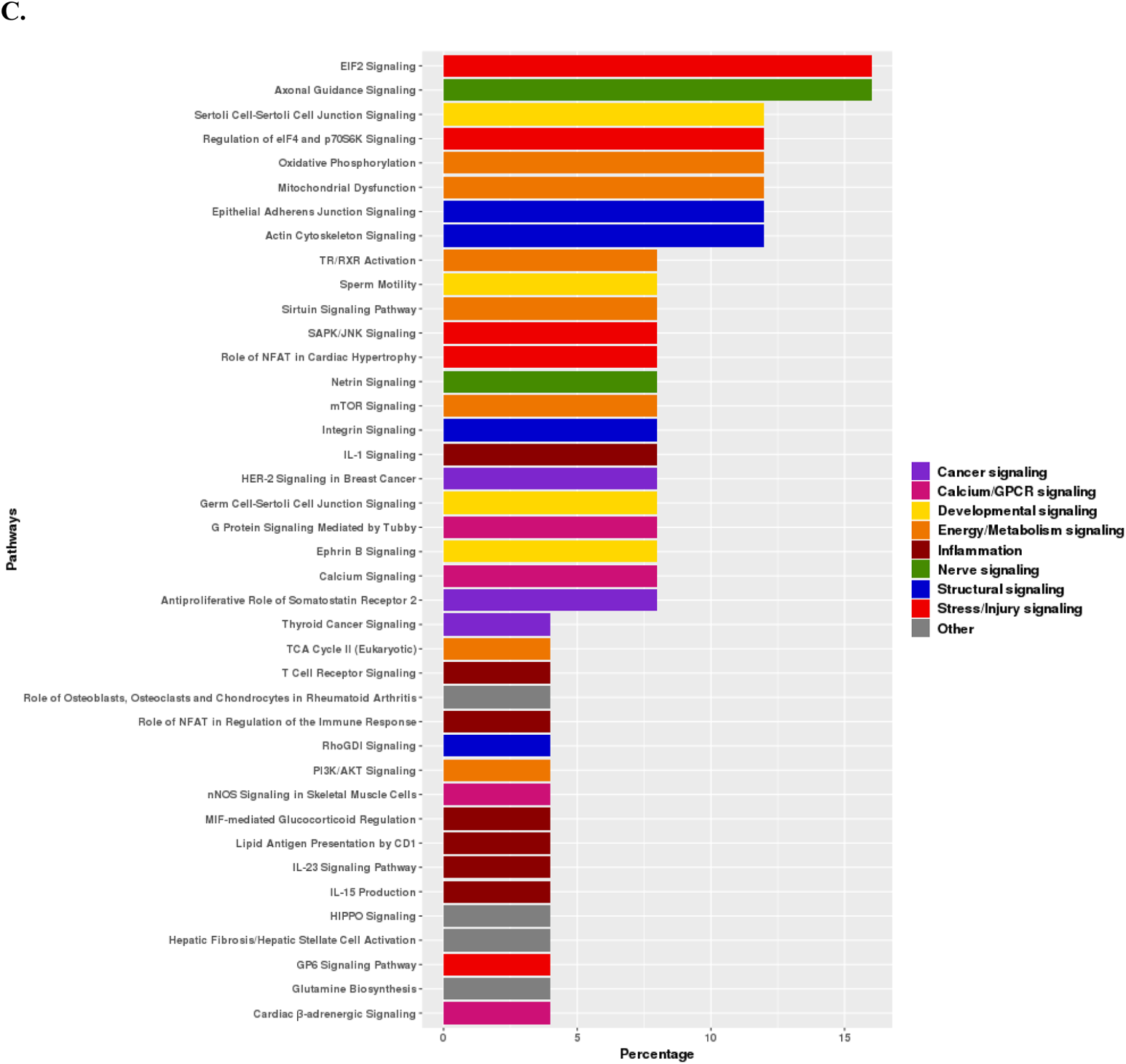
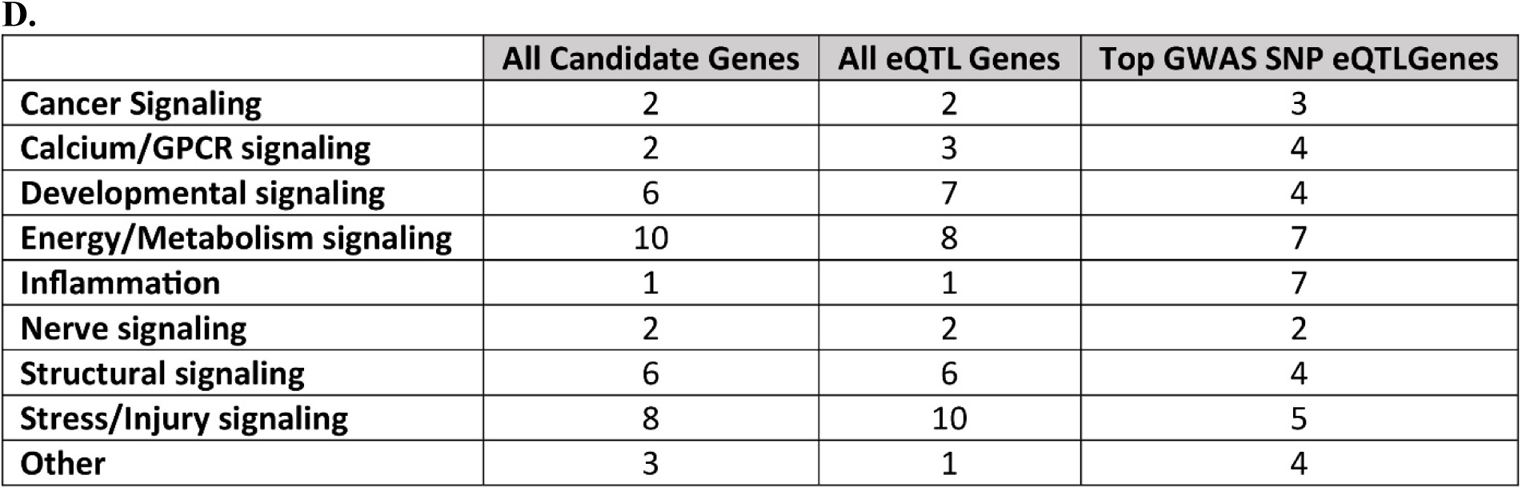
Top significant IPA® canonical pathways for the coexpression networks of each candidate AF gene list. IPA canonical pathways for each candidate gene were ranked from lowest to highest p-value and the top 5 significant canonical pathways listed for each gene (See Supplemental Excel file 8, Supplemental Figure 3-5). Displayed here is the percentage of genes for which canonical pathways were listed in the top 5. Data are displayed by gene lists. **A. All Candidate Genes** (N=225; data not available for 8 genes, see text). Mitochondrial dysfunction, oxidative phosphorylation, epithelial adherens junction signaling, sirtuin signaling pathway and actin cytoskeleton pathway were the most frequent pathways listed among the top 5 canonical pathways. **B. Any eQTL Genes** (n=131). Epithelial adherens junction signaling, mitochondrial dysfunction and oxidative phosphorylation, germ cell-Sertoli cell junction singnaling and axonal guidance signaling were the most frequent pathways listed among the top 5 canonical pathways. **C. Top GWAS SNP eQTL Genes** (n=37). EIF2 signaling, axonal guidance signaling, regulation of eIF4 and p70S6K signaling, oxidative phosphorylation, and mitochondrial dysfunction were the most frequent pathways listed among the top 5 canonical pathways. **D. Frequency of significance of each pathway for the 3 gene lists.** Energy/Metabolism signaling followed by Stress/Injury signaling, followed by Developmental signaling and Structural signaling were the most frequent themes in the All Candidate Genes list. Stress/Injury signaling, followed by Energy/Metabolism signaling, Developmental signaling, and Structural signaling were the most frequent themes in the All eQTL Genes list. Energy/Metabolism signaling and Inflammation most commonly appeared in the Top GWAS SNP eQTL Genes list.

**Supplemental Table 1.** Top pathways and upstream regulators within each AF candidate gene list.

|  | Top GWAS SNP eQTL Genes (n=37) | All eQTL Genes (n=131) | All Candidate Genes (n=233) |
| --- | --- | --- | --- |
| Top Canonical Pathways | • G Beta Gamma Signaling<br>• Ceramide Degradation<br>• Gi Signaling<br>• Sphingosine and Sphingosine -1-phosphate Metabolism<br>• Axonal Guidance Signaling | • IL-15 Production<br>• G Beta Gamma Signaling<br>• Caveolar-mediated Endocytosis Signaling<br>• Phagosome Maturation<br>• Agrin Interactions at Neuromuscular Junction | • Cardiac Hypertrophy Signaling (Enhanced)<br>• Axonal Guidance Signaling<br>• Cardiomyocyte Differentiation via BMP Receptors<br>• Ephrin Receptor Signaling<br>• Factors Promoting Cardiogenesis in Vertebrates |
| Top upstream regulators | • miR-4455<br>• *MEF2C*<br>• *PTCH1*<br>• *HEY2* | • *ESRRA*<br>• miR-450a-1-3p<br>• *LRIG3*<br>• Phenylacetate | • *MYOCD*<br>• *ESRRA*<br>• *NKX2-5*<br>• *HEY2*<br>• *MEF2C* |
| Top Networks | • Cancer, Organismal Injury and Abnormalities, Respiratory Disease<br>• Organ Morphology, Organismal Injury and Abnormalities, Skeletal and Muscular Disorders | • Developmental Disorder, Hereditary Disorder, Organismal Injury and Abnormalities<br>• Cardiac Arrhythmia, Cardiovascular Disease, Organismal Injury and Abnormalities<br>• Auditory Disease, Cardiovascular Disease, Dental Disease<br>• Organismal Injury and Abnormalities, Skeletal and Muscular Disorders, Infectious Diseases<br>• Cancer, Organismal Injury and Abnormalities, Respiratory Disease | • Cardiovascular Disease, Organismal Injury and Abnormalities, Skeletal and Muscular Disorders<br>• Cell-To-Cell Signaling and Interaction, Cellular Assembly and Organization, Cancer<br>• Cardiovascular Disease, Hereditary Disorder, Organismal Injury and Abnormalities<br>• Inflammatory Disease, Infectious Diseases, Lipid Metabolism<br>• Cell Cycle, Developmental Disorder, Hereditary Disorder |

